# Building an AI-Powered Educational Tool for Exploring Microbial Relationships in Parkinson’s Disease

**DOI:** 10.1101/2025.09.18.25335721

**Authors:** Sophie Shih, Hyunji Kim, Young-tak Kim, Synho Do

**Affiliations:** Mass General Brigham

## Abstract

This paper presents the Neurobiome Navigator, an AI-powered, highly interactive, and easily navigable application designed to help users explore the complex relationships between the human microbiome and Parkinson’s disease (PD). The app focuses on the gut microbiome and the oral microbiome, known to have a strong relationship with PD, as well as impulse control disorders (ICD), a significant non-motor symptom of PD. The system integrates the MINERVA (Microbiome Network Research and Visualization Atlas) knowledge graph with supplemental scientific literature to deliver evidence-based insights. Built using an AI agentic workflow including Streamlit, Neo4j, and LangChain, the application enables users to submit structured survey responses (e.g., oral health, impulse control) or free-text queries. Data retrieval methods include Neo4j graph queries and semantic vector search. Retrieved content is synthesized by a large language model (GPT-4o) and an agentic pipeline to generate personalized, non-clinical suggestions. By bridging advanced AI capabilities with an accessible interface, the Neurobiome Navigator aims to empower users to make informed lifestyle decisions that can potentially improve quality of life, particularly in managing non-motor symptoms of PD. The platform enhances understanding of microbiome-PD connections by presenting complex scientific relationships in an interactive, visually engaging, and easily navigable format. Through personalized insights, dynamic charts, and exploratory tools, it transforms dense biomedical data into actionable, user-friendly guidance, making the learning process both informative and enjoyable.

## 1. Introduction

Parkinson’s Disease (PD) is a progressive neurodegenerative disorder characterized by motor symptoms such as tremors and rigidity, as well as non-motor symptoms including impulse control disorders (ICD) and sleep disturbances. Increasing evidence suggests that imbalances in microbiota, termed dysbiosis, play a major role in PD onsight and progression. Gut and oral dysbiosis have emerged as parallel contributors, potentially influencing the disease through similar inflammatory and neurological pathways [1].

Despite rapid advances in microbiome research, knowledge remains scattered across the scientific literature, hindering a comprehensive understanding of the current state of the field and the intricate web of microbiome influences on human health. Variations in study populations, analytical techniques, and data processing approaches further contribute to inconsistencies in findings, making it difficult for researchers, clinicians, and students to synthesize information into a coherent, reliable picture. These challenges are reflected in PD research, where variability in study methods, population characteristics, and the heterogeneity of PD itself contribute to inconsistent findings. Still, multiple studies have identified recurring microbial patterns such as reduced short-chain fatty acid-producing bacteria and increased pro-inflammatory taxa [2], supporting the microbiome’s potential role as both a biomarker and a therapeutic target.

At the core of this thesis is the Neurobiome Navigator, an AI-powered educational application developed to help users explore the complex and fragmented research connecting PD to the human microbiome. Designed to be accessible to both specialists and non-specialists, the Neurobiome Navigator focuses on three key areas of emerging interest: gut dysbiosis, oral dysbiosis, and ICD. Its goal is to transform highly technical, dispersed findings into an interactive, evidence-based resource that supports exploration and understanding without requiring deep domain expertise. Beyond exploration, the goal of the app is to deepen understanding and support informed, lifestyle-based decisions that may contribute to risk reduction.

The application is built on AI, particularly large language models (LLMs) integrated with knowledge graph technology. It draws heavily on MINERVA [3] (Microbiome Network Research and Visualization Atlas), a robust, evidence-linked knowledge graph mapping over 66,000 microbe–disease associations from more than 129,000 publications. By using MINERVA as a structured backbone and supplementing it with additional scientific literature, the Neurobiome Navigator synthesizes information from multiple trusted sources into a unified, coherent platform.

Through conversational AI, engaging visuals, and the ability to connect personal input to research-backed insights, the Neurobiome Navigator makes it possible to navigate microbiome–PD research in an approachable yet scientifically rigorous way. In doing so, it bridges the gap between vast, unstructured scientific knowledge and the accessible, interpretable formats needed for education, hypothesis generation, and broader scientific engagement.

## 2. Background & Related Work

### 2.1 Parkinson’s and the Gut-Brain Axis

PD is a prevalent neurodegenerative disease characterized by the aggregation of alpha-synuclein (α-syn), a neuronal protein involved in the regulation of synaptic vesicle trafficking and the release of neurotransmitters, and the progressive degeneration of dopaminergic neurons within the substantia nigra. Beyond its well-known motor symptoms like bradykinesia, slowness in movement or speed; resting tremor; and rigidity, PD also presents significant non-motor symptoms, such as ICD, gastrointestinal dysfunction and sleep disturbances, all of which can severely impair a patient’s quality of life. Currently, treatment for PD relies on pharmacotherapy and surgical interventions to manage symptoms, but effective disease-modifying therapies that can reverse α-syn aggregation or restore dopaminergic neuron degeneration are still lacking [4].

Recent research has introduced a new perspective, highlighting the potential impact of the gut microbiota on the central nervous system. The gut microbiota refers to the complex community of bacteria, viruses, fungi, and other microorganisms that inhabit the intestinal tract, representing the largest and most intricate microflora in the human body. The “gut microbiota-gut-brain axis” (MGBA) is a complex bidirectional communication pathway linking the gut microbiota, the gut, and the brain. It enables the enteric nervous system (ENS) and the central nervous system (CNS) to communicate through neurological, immune, hormonal, and metabolic signals. Research increasingly shows that gut dysbiosis may play a key role in PD, influencing its onset, severity, and progression. Mechanistically, it may contribute through increased intestinal permeability (“leaky gut”), inflammation in the gut and brain, abnormal α-syn aggregation, oxidative stress, and reduced neurotransmitter production. One key hypothesis suggests PD may begin in the gut and spread to the brain via the vagus nerve, making the gut microbiota a promising target for treatment [4].

ICDs in PD, traditionally linked to dopaminergic therapy, have also been associated with gut microbiota dysbiosis. Specific bacterial taxa, such as Methanobrevibacter and Intestinimonas butyriciproducens, along with altered microbial metabolic pathways, may influence impulse control through effects on neurotransmitter production, neuroinflammation, and brain function [2]. In addition, oral dysbiosis—an imbalance in the oral microbiome—has been linked to systemic inflammation, neurodegeneration, and symptom progression through the oral–gut–brain axis. These microbial changes, often driven by periodontal disease, may influence PD pathogenesis by promoting neuroinflammation, disrupting the blood–brain barrier. The oral microbiome can serve as another potential biomarker for early diagnosis and disease monitoring [1].

### 2.2 Microbiome Complexity: Variables and Species Involved

The human microbiome is an extremely complex biological system. As the largest and most intricate microflora in the human body, it encompasses approximately 50 bacterial phyla. Under normal conditions, the abundance and diversity of the gut microbiota and oral microbiota are in a dynamic balance. However, factors such as diet, stress, and antibiotics can profoundly impact the gut microbial community structure, and can have similar effects on the oral microbial community structure. Dysbiosis is involved in the development of many conditions, including intestinal diseases (e.g., inflammatory bowel disease, irritable bowel syndrome), mental disorders (e.g., anxiety, depression, autism spectrum disorder), and neurological diseases (e.g., multiple sclerosis, Alzheimer’s, Parkinson’s, and amyotrophic lateral sclerosis) [5].

Microbiome research has grown rapidly, with over 129,000 publications between 2014 and 2023 alone, making it difficult to synthesize such vast, complex data. Variations in populations, methods, and analysis can produce inconsistent results, requiring researchers to consult multiple sources. Manual curation has created valuable databases like HMDAD, gutMDisorder, and Disbiome, but these struggle to keep pace with the field’s growth and are limited in providing insights beyond basic data retrieval [3].

### 2.3 Knowledge Graphs and AI Tools in Biomedicine: The Role of MINERVA and AI

In biomedicine, knowledge graphs (KGs) organize complex, scattered information into networks of interconnected entities (nodes) and relationships (edges). This structure is especially valuable in microbiome research, where data spans thousands of studies. MINERVA (MIcrobiome NEtwork Research and Visualization Atlas) exemplifies the power of AI-powered KGs, mapping microbe–disease associations from scientific literature using a fine-tuned Large Language Model. It enriches each connection with metadata, evidence links, and relevance scores, creating an interactive, evidence-grounded resource. To address LLM “hallucinations,” MINERVA cross-checks findings across multiple sources and links every claim directly to supporting text, ensuring accuracy and transparency [3].

AI frameworks such as LangChain [6] allow for the integration of LLMs with databases and retrieval systems, while vector search allows precise matching of user queries to relevant scientific texts. Paired with an intuitive frontend, these tools make vast biomedical datasets, more accessible and easier to explore for users. By leveraging AI, static knowledge can be turned into a dynamic, searchable, and user-friendly resource.

## 3. System Design & Implementation

This project focuses on building a prototype that bridges complex research with a user-friendly interface, using recent AI tools. While platforms like MINERVA excel at extracting and structuring complex biomedical knowledge, their capabilities can be difficult for non-experts to navigate. By combining robust data processing pipelines with intuitive visualizations and guided exploration features, the prototype transforms dense scientific resources into an engaging, evidence-grounded decision-support tool for navigating PD and microbiome relationships.

### 3.1 System Architecture Overview

**Figure 3.1:**
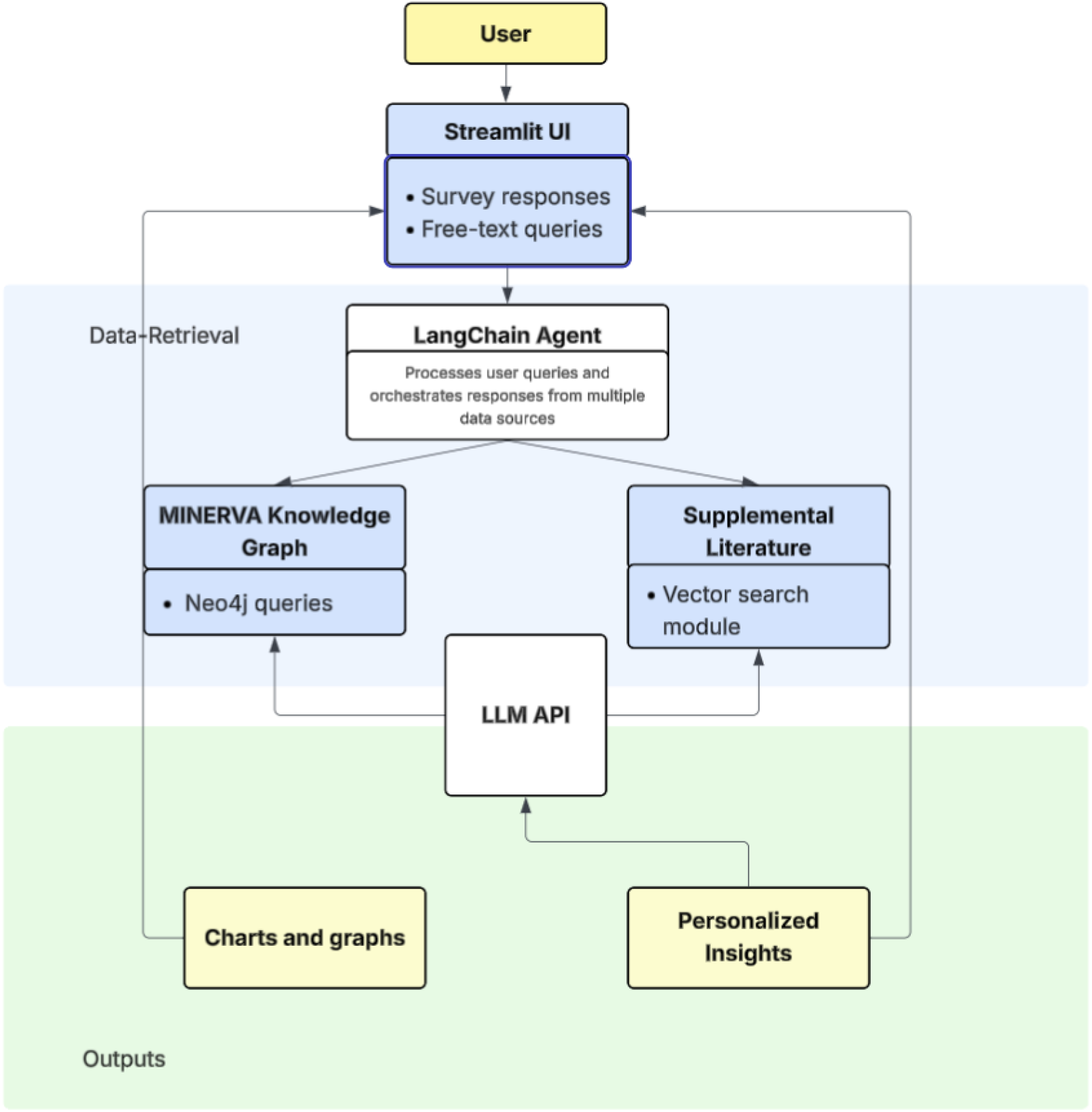
System architecture of the Neurobiome Navigator application, illustrating the flow from user interaction in the Streamlit UI through application logic, data retrieval, and output generation.

### 3.2 Technologies Used

The Neurobiome Navigator was created by a system that integrates a wide range of graph database technologies, web frameworks, and AI tools. At its core, the application connects to MINERVA’s Neo4j graph database, which stores large-scale, biomedical associations derived from literature. Neo4j was used as the primary graph database due to its ability to dynamically handle complex relationships between microbiome and disease entities [7]. The frontend was developed using Streamlit [8], a lightweight web application framework suited for quickly building and deploying powerful data applications. Custom CSS and HTML were used to enhance layout and styling.

A custom AI agent was developed using the Pydantic AI [9] framework to interpret natural language queries, retrieve relevant biomedical data, and allows for user-specific questions. This agent was built using LangChain [6], which enabled conversational memory and tool integration, and the OpenAI API [10], which provided natural language processing and generation capabilities. To handle the semantic search aspect of the application, FAISS [11] was used for vector similarity search, particularly for indexing and querying research papers. PyMuPDF (fitz) [12] handled PDF parsing and text extraction. The combination of semantic searching and querying the MINERVA graph database allowed the system to incorporate external literature to provide richer, evidence-based responses.

This development process was supported by Docker [13], which ensured a consistent runtime environment across machines, and Docker Compose [14], which coordinated multiple service containers for Streamlit and Neo4j.

### 3.3 Key Features

A central feature of the application is its natural language querying capability, which enables users to interact with the MINERVA knowledge graph and supplemental literature sources without requiring technical query syntax. This allows flexible exploration of complex biomedical relationships for non-expert users.

The system also incorporates personalized surveys on oral health and impulse control. Upon completing surveys, users are presented with a visual graph that displays their answers. Responses are stored and reflected in a personal snapshot, where users can see a tailored profile summarizing their survey answers and general lifestyle-based recommendations. To enhance relatability, the application includes sample snapshots representing realistic PD scenarios at varying stages.

Dedicated insight pages allow users to focus on a specific domain of interest. The Microbiome Insights page presents information on individual microbiomes known to influence target areas such as ICD, gut health, and oral health. The Food Insights page draws on MINERVA’s food-microbiome relationship data to provide detailed microbial information for selected from a list of the top five protective and harmful foods associated with PD. The Research Insights page allows users to submit custom questions, which are answered using the semantic search pipeline over scientific literature.

### 3.4 Development Approach

This application was primarily developed through an iterative prototyping process, also known as vibe coding, using the Windsurf [15] development environment. Rather than following a fully test-driven approach, the focus was on rapid experimentation and incremental refinement. This allowed for features such as personalized surveys and AI-driven queries to be tested quickly with real data from MINERVA’s Neo4j graph. Throughout development, usability was prioritized over full backend mastery. The goal was to produce a functional and intuitive interface, even if certain backend components were implemented in minimal form.

## 4. Demonstration & Results

### 4.1 Use Case Scenarios

To demonstrate the functionality and educational value of the application, two representative use cases are presented. Each scenario shows how a user might explore specific microbiome–disease or food–microbiome relationships using the interactive interface. Screenshots are included to illustrate the workflow and outputs.

#### Use Case 1: Exploring Lachnospiraceae’s role in PD

**Figure 4.1:**
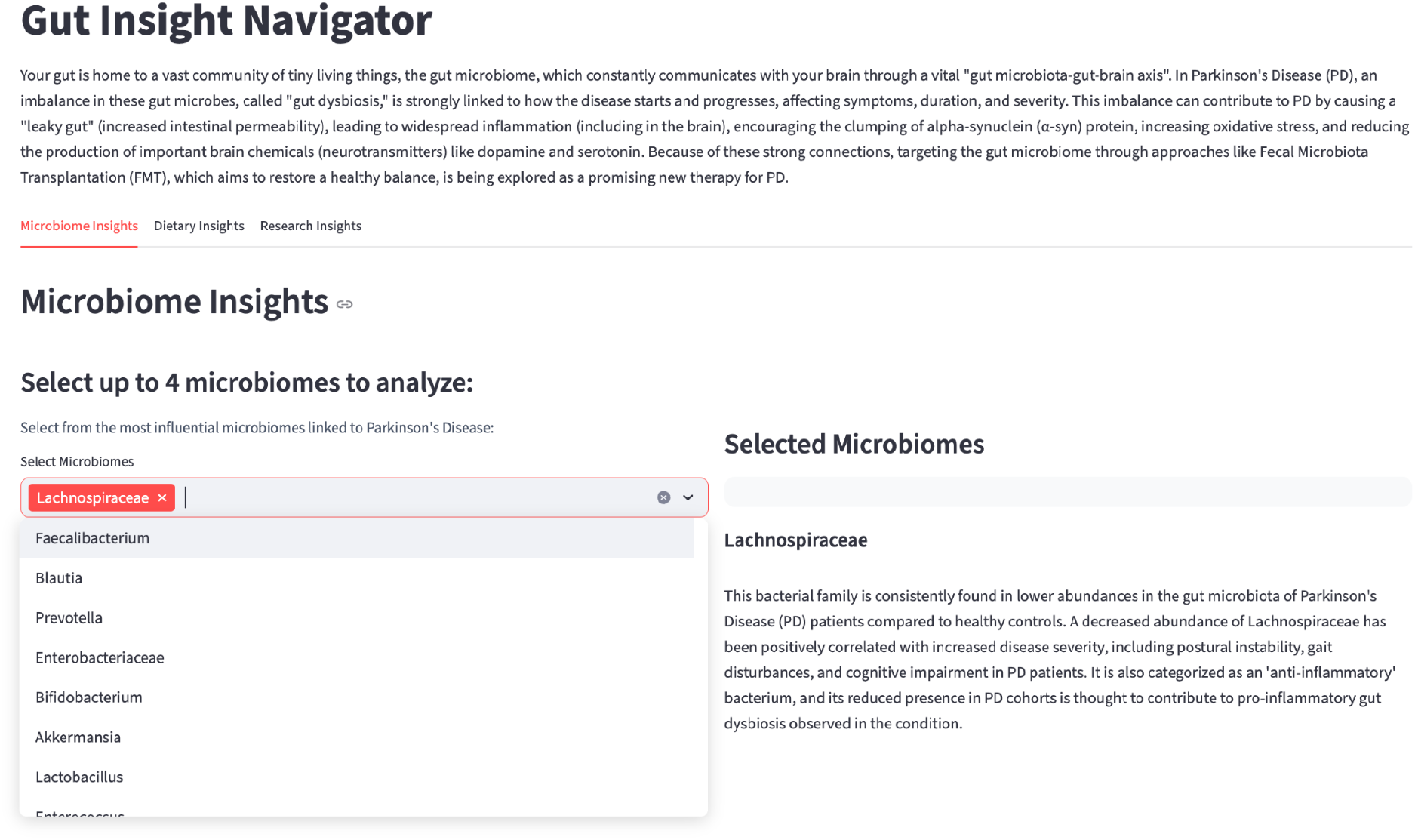
Microbiome Insights page of the Gut Insight Navigator showing Lachnospiraceae selected for analysis. The right-hand panel displays a literature-derived description highlighting its reduced abundance in PD, its anti-inflammatory role, and its correlation with symptom severity.

In the Microbiome Insights page of the Gut Insight Navigator, the user selects “Lachnospiraceae” from the list of available microbiomes. The application displays a detailed description derived from recent literature, highlighting Lachnospiraceae’s reduced abundance in PD and its role as an anti-inflammatory bacterium. This view enables the user to connect specific microbiome shifts to disease symptoms and progression.

#### Use Case 2: Investigating food-microbiome relationships in the context of PD

**Figure 4.2a:**
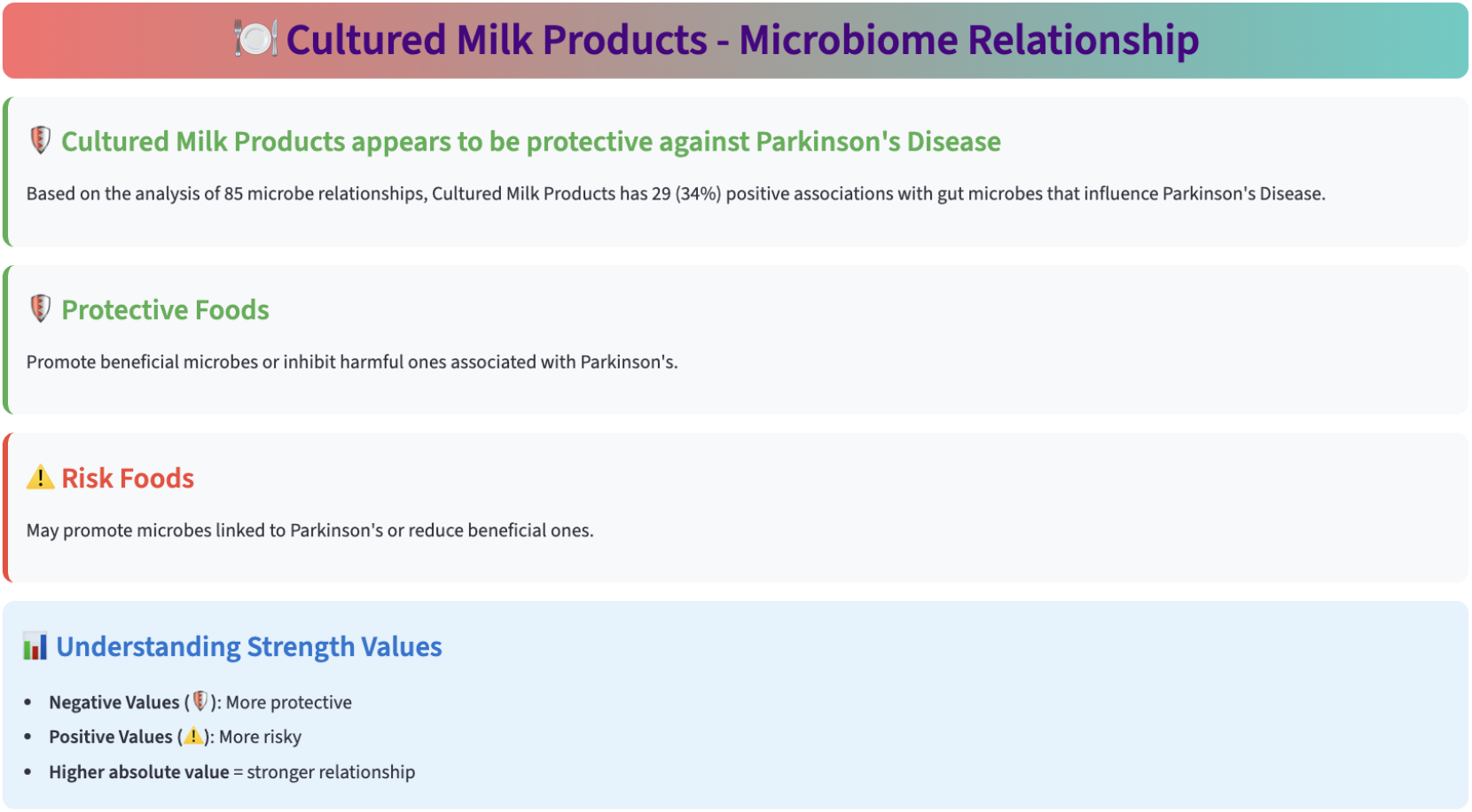
Food Insights view for the selection, “cultured milk products”. This section provides an overview of protective vs. risk classification and visual key for interpreting relationship strength values.

**Figure 4.2b:**
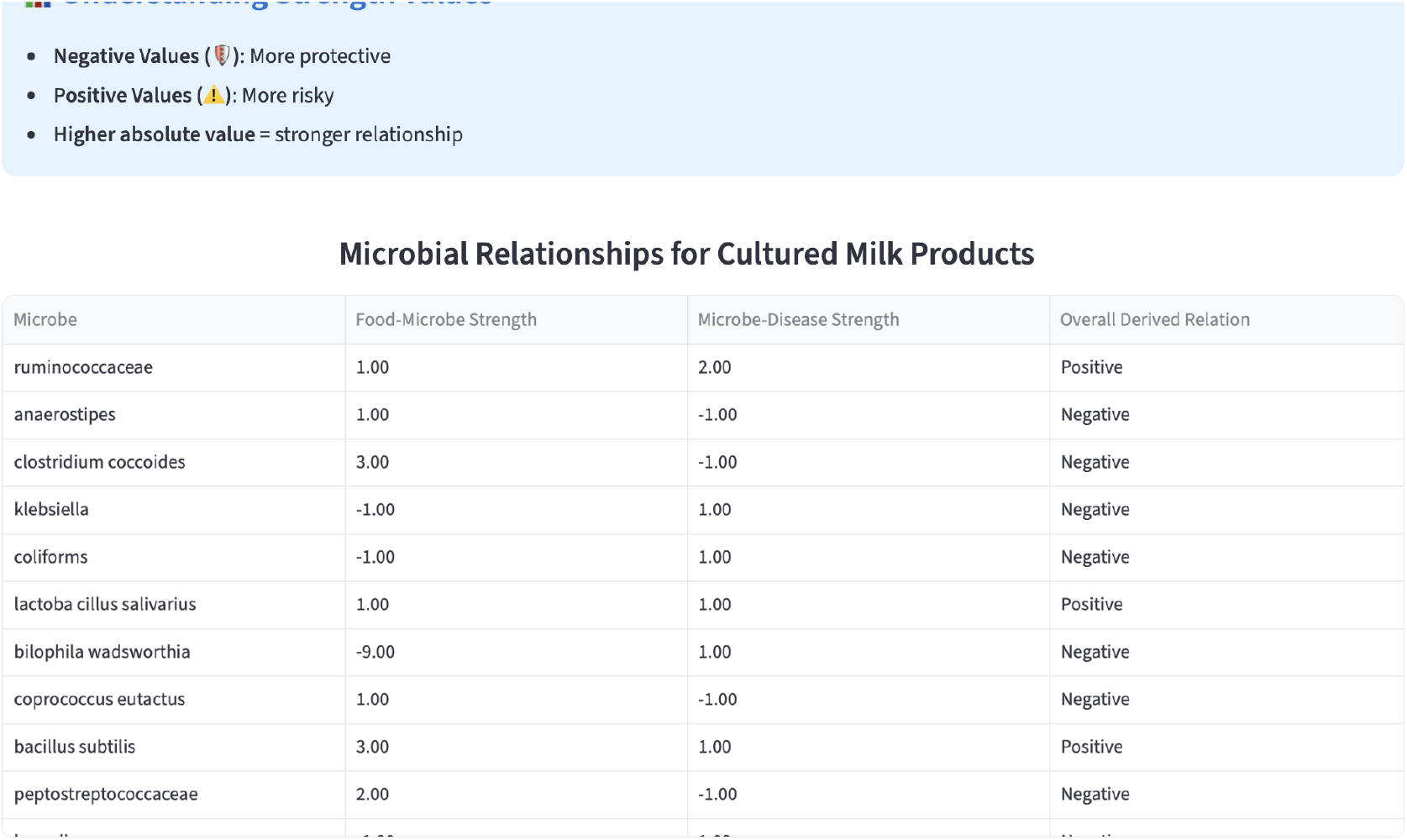
Microbial relationship table for cultured milk products, showing the strength and direction of associations between the food item, individual microbes, and PD.

In the Food Insights tab, the user selects “Cultured milk products” from a list of the top five protective foods against PD to explore potential microbiome impacts. The application displays a color-coded explanation of “protective” versus “risk” foods and includes a visual key for interpreting relationship strength values. Below this, a table lists relevant microbes affected by the selected food, along with relationship types and confidence levels, drawn from MINERVA. This structure enables both quick visual interpretation and a detailed breakdown in table form.

### 4.2 User Experience & Output Quality

Overall, the platform is designed to be accessible to non-technical users, including patients and general audiences. While some microbiome descriptions are dense with scientific terms and the names of microbiome species, potentially making them more challenging for certain users to read, most elements are written with readability in mind. The application offers unique value by providing microbiome-level insights that are often absent from mainstream health resources, along with connections to dietary patterns, impulse control, oral health, and everyday lifestyle factors that are relevant to PD.

The interface emphasizes a colorful, engaging visual style, particularly on the home screen, the Food Insights page, and within the sample snapshot pages. Navigation is designed to be intuitive, with clear options for selecting specific microbes, completing personalized surveys, and exploring food–microbiome relationships. These interactive elements support both exploratory learning and targeted information retrieval, making the experience appealing for a wide range of users.

The application consistently returns information grounded in the MINERVA knowledge graph and a curated set of supplemental sources. While certain outputs such as personalized insights generated from survey responses are dynamically produced by the AI agent at runtime, many of the microbiome-specific insights are intentionally hard-coded. This ensures that core content, such as the microbiome descriptions, remains accurate, relevant, and consistent across sessions. These hard-coded elements are derived directly from recent literature and MINERVA data, with users given the option to display or hide them for a more interactive and customizable experience.

### 4.3 Performance

**Table 4.3.** Summary of recorded performance metrics for ICD and Oral Health surveys, showing average, minimum, and maximum completion times in milliseconds for chart rendering and LLM insight generation.

| Metric | Avg Time (ms) | Min Time (ms) | Max Time (ms) | Count |
| --- | --- | --- | --- | --- |
| insights_chart_render | 15.20 | 5.40 | 61.77 | 7 |
| insights_llm_generation | 38,171.56 | 27,289.64 | 49,053.47 | 6 |

Two key performance metrics were recorded for the ICD and Oral Health survey workflows: insights_chart_render and insights_llm_generation. insights_chart_render measures the time required to produce the visual charts once a user submits their survey responses. This process was consistently fast, averaging 15.20 ms with a minimum of 5.40 ms and a maximum of 61.77 ms, indicating highly responsive UI updates and minimal impact on the overall user experience.

insights_llm_generation captures the time taken for the AI model to generate personalized insights based on the survey responses. This step was significantly slower, with an average completion time of ~38 seconds, a minimum of ~27 seconds, and a maximum of ~49 seconds. The long duration is mainly due to delays from external API calls and network latency. Since this step takes far longer than chart rendering, it should be the focus for optimization.

insights_llm_generation captures the time taken for the AI model to generate personalized insights based on the survey responses. These insights are produced by an AI agent specifically tailored to interpret and respond to each individual’s survey profile, integrating results from the MINERVA knowledge graph and supplemental literature. This step was significantly slower, with an average completion time of ~38 seconds, a minimum of ~27 seconds, and a maximum of ~49 seconds. The extended duration is largely due to delays from external API calls and network latency, compounded by the additional processing required to synthesize tailored, context-specific recommendations. Since this stage takes far longer than chart rendering, it should be prioritized for optimization.

## 5. Discussion

### 5.1 Strengths of the Application

One strength of the application is its ability to make the complex biomedical relationships linked to PD explorable without requiring deep technical expertise. By abstracting away the intricacies of database querying, the system allows users to investigate the role of specific microbiomes, dietary factors, and lifestyle elements in PD without needing prior experience with tools like Neo4j.

The platform also demonstrates the potential of combining AI with biomedical knowledge graphs in a PD-focused setting. By integrating an AI agent, semantic search capabilities and the MINERVA Neo4j knowledge graph, the application displays microbiomes, symptoms, and environmental factors relevant to PD. This not only highlights potential areas to further research but also shows how AI-driven exploration can accelerate the understanding of complex conditions.

In addition, the system offers significant educational potential for audiences seeking to better understand PD. Students can use it to learn about the underlying biology and research landscape; clinicians can explore emerging relationships between microbiome changes and disease progression; and patients or caregivers can gain insight into how dietary and lifestyle factors intersect with the condition. By combining personalization, interactivity, and a research-backed knowledge base, the prototype fosters informed engagement with a complex and evolving area of study.

### 5.2 Limitations

While the prototype demonstrates the potential of combining AI and biomedical knowledge graphs, several limitations should be acknowledged. First, the system relies on LLM-driven query generation and abstraction through LangChain, meaning much of the underlying query construction was handled automatically during development. While this approach accelerated prototyping and reduced the need for manual query design, it also limited transparency into the exact query mechanics, making it harder to verify or fine-tune.

Second, the biomedical accuracy of the presented insights has not been validated by domain experts. Although the application draws from the reputable MINERVA knowledge graph and supplements it with semantic search over research literature from PubMed, the findings should be interpreted as exploratory rather than definitive, especially in clinical contexts.

Third, the semantic search component currently operates on a limited supplemental literature set, which restricts the breadth of contextual evidence the system can provide, especially when it comes to custom questions. Combined with the fact that the application covers only a narrow subset of the microbiome–PD research space, focusing primarily on impulse control disorders, gut health, and oral health, this means that many relevant microbiome–disease relationships remain outside the scope of the current version. Furthermore, given the vast diversity of microbes potentially relevant to PD, the application currently focuses in detail on only a select group of microbiomes known from literature to be among the most influential. Many of these details are hardcoded into the application’s logic, meaning that expanding the scope to new microbiomes would require manual updates, making large-scale scaling inefficient in its current form.

Finally, the prototype remains in an early-stage form, with opportunities to enhance both the user interface and the depth of query results. Future refinements could include richer visualizations, expanded exploration tools, response-time optimization, and a more comprehensive dataset integration to capture the complexity of PD-microbiome research.

### 5.3 Future Work

Future development will focus on expanding the system’s scope, improving accuracy, and enhancing user engagement. A key next step is to gather and incorporate user feedback and conduct targeted testing with PD patients, caregivers, and clinicians. This will help refine both the interface and the types of insights presented, ensuring the platform is aligned with real-world needs and expectations.

From a data perspective, the application could benefit from deeper integration with biomedical resources such as PubMed and specialized microbiome datasets. This would expand the coverage beyond the current select set of influential microbes, providing a more comprehensive view of microbiome–PD relationships.

From a data perspective, the application could benefit from deeper integration with biomedical resources such as PubMed and specialized microbiome datasets. This would expand coverage beyond the current select set of influential microbes, providing a more comprehensive view of microbiome–PD relationships. To make scaling more efficient, the application architecture could be adapted to more dynamically ingest and process new data rather than relying on hardcoded microbiome details. This system would allow for the automatic incorporation of new studies, microbes, and disease associations, enabling the app to evolve with the research landscape.

Finally, the visualization layer could be enhanced with more dynamic and interactive elements, such as real-time rendering of nodes and edges directly from the knowledge graph. This would allow users to trace connections between microbes, dietary factors, and symptoms in a more intuitive and exploratory way, deepening their engagement with the underlying data.

## Conclusion

This project set out to confront a central challenge in PD research: the difficulty of synthesizing a growing yet fragmented body of evidence linking the disease to gut and oral dysbiosis, as well as ICD. The objective was to develop an AI-powered educational application capable of uniting these scattered findings into an accessible, interactive resource. Built on the foundation of MINERVA’s evidence-linked knowledge graph, the Neurobiome Navigator enables users to explore microbial associations with PD, across gut dysbiosis, oral dysbiosis, and ICD, through natural language queries, engaging visuals, and integration of personal survey data. This app facilitates exploration and understanding even for those without domain-specific expertise. Its design emphasizes interpretability and engagement, allowing patients, students, researchers, and clinicians alike to navigate complex biomedical relationships in a way that is both grounded in evidence and intuitively presented.

Beyond its immediate functionality, this project highlights the broader potential of AI-powered tools to democratize access to specialized scientific insight. By translating technically demanding microbiome-PD research into a coherent and navigable form, it offers a model for how AI can bridge the gap between complex, disparate scientific data and the diverse audiences who stand to benefit from it.

## Data Availability

All data produced in the present work are contained in the manuscript

Neurobiome Navigator: https://neurobiomenavigator.mllm.bet/

Neurobiome Navigator Walkthrough: https://www.youtube.com/watch?v=CgUKoGj21uI

Neurobiome Navigator Github: https://github.com/sophies60/Neurobiome-Navigator/tree/main

